# Association between Self-reported Masking Behavior and SARS-CoV-2 Infection Wanes from Pre-Delta to Omicron-Predominant Periods — North Carolina COVID-19 Community Research Partnership

**DOI:** 10.1101/2022.05.27.22275689

**Authors:** Ashley H. Tjaden, Michael Gibbs, Michael Runyon, William S. Weintraub, Yhenneko J. Taylor, Sharon L. Edelstein, the COVID-19 Community Research Partnership Study Group

**Affiliations:** Biostatistics Center, Milken Institute School of Public Health, George Washington University; Department of Emergency Medicine, Atrium Health; MedStar Health Research Institute and Georgetown University; Center for Outcomes Research and Evaluation, Atrium Health

**Keywords:** COVID-19, mask use

## Abstract

We assessed the association between self-reported mask use during non-household interactions and COVID-19 infection during three pandemic periods. Odds of infection for those who did not always compared to those who always wore a mask was 66% higher during pre-Delta, 53% higher during Delta, declining to 16% higher during Omicron.

## Introduction

Wearing a facemask has been a primary public health method to reduce SARS-CoV-2 transmission throughout the pandemic.^1,2,3^ However, how well masks conveyed protection during each variant wave, including the most recent Omicron wave, has yet to be evaluated. Using prospectively collected data from the North Carolina COVID-19 Community Research Partnership (CCRP), we assessed the association between self-reported mask use when interacting with others outside the household and risk of COVID-19 infection during three periods of the pandemic.

## Methods

The NC CCRP^4^ is a prospective, multi-site cohort COVID-19 syndromic surveillance study of a convenience sample of participants enrolled primarily through direct email outreach at six NC healthcare systems from April 2020 through June 2021 (http://www.covid19communitystudy.org/). Data were collected via a secure, HIPAA-compliant, online platform through March 2022. All participants provided informed consent, and Institutional Review Board (IRB) approval was provided by the Wake Forest School of Medicine. We performed a nested case-control analysis within the study cohort to compare self-reported cases to controls who never self-reported a positive test for SARS-CoV-2 infection. For this report, eligible participants were ≥18 years, completed daily syndromic surveillance surveys, did not participate in a vaccine trial, and did not self-report prior COVID-19 infection at enrollment (**Supplemental Figure 1**). Demographic data and healthcare worker status were collected at enrollment. Daily online email or text surveys asked about COVID-19 symptoms, test results, vaccination, and risk behavior. Not wearing a mask was defined as responding “no” at least once in the ten days preceding the match date to the daily survey question, “In the last 24 hours, have you worn a face mask or face covering every time you interacted with others (not in your household) within a distance of less than 6 feet?” We defined SARS-CoV-2 vaccination using participant self-report and categorized participants into three categories: at least 14 days after receiving second of two doses of either the Pfizer BioNTech BNT162b2 or Moderna mRNA-1273 vaccine, receiving at least one dose of either mRNA vaccine or unvaccinated. Participants who reported receiving a non-mRNA vaccine or had an undetermined vaccine status were excluded.

We matched up to 10 controls to each case on calendar time of first self-reported positive test. Case participants self-reported at least one infection (positive SARS-CoV-2 antigen or nucleic acid amplification test) during study follow-up. To address multiple eligible survey entries available for matching per control, we used an optimal matching algorithm maximizing the cases included in the analysis.^5^ The number of controls per case ranged from 1 to 10 with a median of 7. Descriptive statistics are presented as absolute and relative frequencies with p-values from unadjusted Pearson’s chi-squared tests and standardized mean differences. Conditional logistic regression models of COVID-19 infection were adjusted for enrollment site, age group, race/ethnicity, county population density (urban/suburban/rural), healthcare worker occupation, vaccination status, and recent known exposure to COVID-19. Covariates in the multivariable model were selected a-priori. After assessing whether there was an interaction between variant predominant period and mask use, we evaluated three periods during the pandemic: Pre-Delta (July 1 2020 – June 30, 2021), Delta (July 1, 2021 – November 30, 2021), and Omicron (December 1, 2021 – February 28, 2022) predominance. Analyses were performed using R (V.4.0.3, R Foundation for Statistical Computing).

## Results

Of the 3,901 cases and 27,813 date-matched controls, participants were majority female (71.5% of cases, 68.6% of controls) and non-Hispanic white (89.1% of cases and 87.5% of controls). Healthcare worker occupation was common (35.4% of cases and 26.8% of controls) (**Supplemental Table 1**). Over half of the cases and controls were matched ≥14 days after a second mRNA vaccine dose (53.7% of cases and 55.7% of controls). About a quarter of participants (25.8% of cases and 27.9% of controls) had at least one comorbidity. The pre-Delta predominant period accounted for 42.5% of cases, compared to 17.1% during the Delta-predominant period and 40.5% during the Omicron-predominant period.

The daily survey response rate in the ten days preceding the match date was 73.1% for cases and 65.3% for controls. Overall, reporting not wearing a facemask when interacting with others outside the household was more prominent among cases prior to the index date (date self-reported positive test) and less prominent after the index date. Approximately 42.5% of cases and 36.3% of controls responded at least once that they did not wear a mask when interacting with others in the 10 days preceding the match date (**Supplemental Table 2**). Mask wearing behavior varied by period in the pandemic (**Figure 1**). Overall, not wearing a mask increased between the pre-Delta and Delta period, then decreasing slightly during the Omicron period. During the pre-Delta period, 28.5% of cases and 19.4% of controls reported at least one day in the 10 days preceding the index date not wearing a mask compared to 61.5% of cases and 55.1% of controls during the Delta period, and 50.2% of cases and 47.5% of controls during the Omicron-predominant period. Over half of the cases (54.1%) reported a known exposure to someone who recently tested positive for COVID-19 in the ten days preceding the index date compared to only 9.3% of controls.

**Figure 1.**
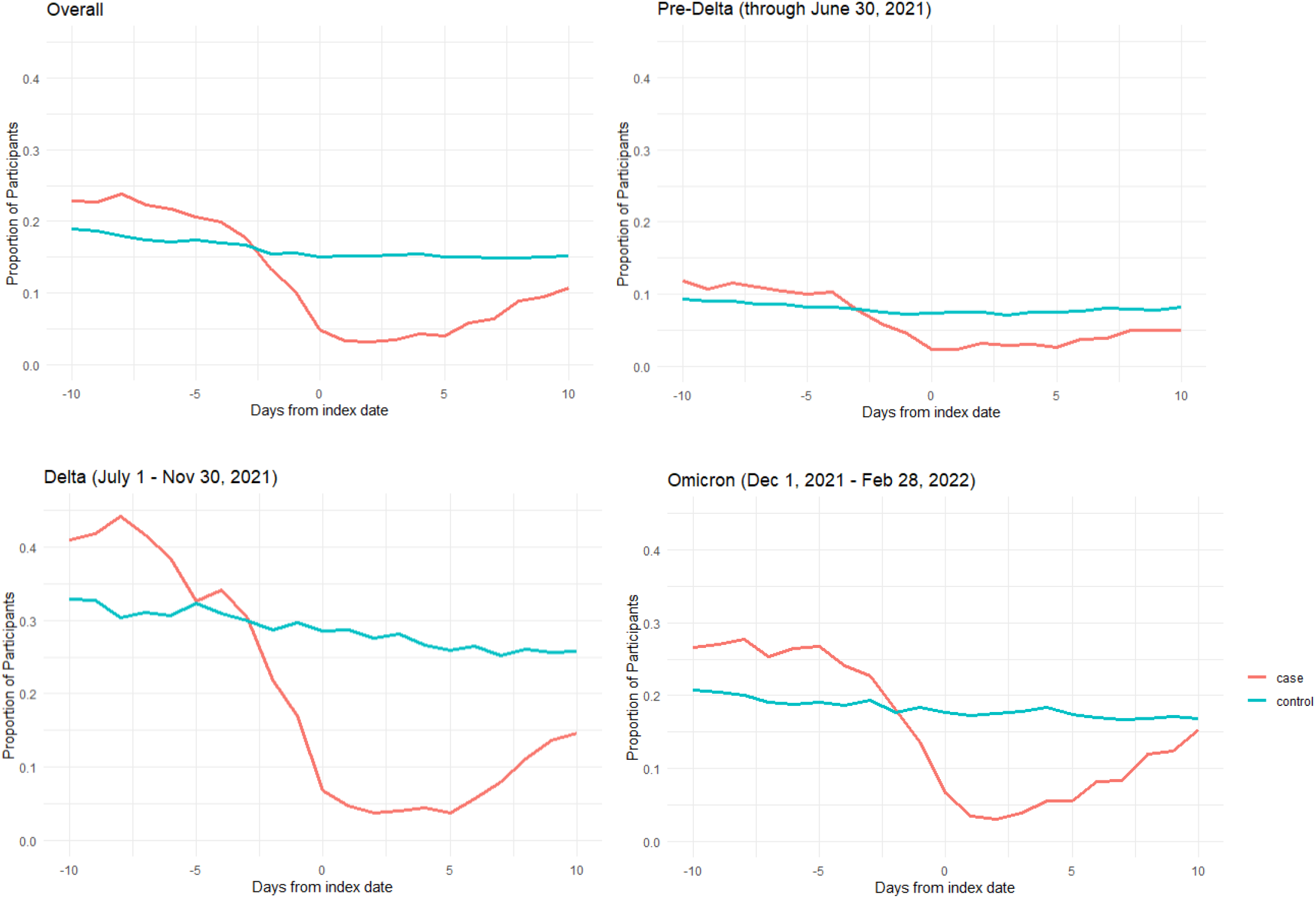
Proportion of Participants Responding to Daily Survey that they did not wear a face mask or face covering every time the interacted with others preceding and following match date (Day 0), by variant-predominant period

The association between mask use and SARS-CoV-2 infection varied by period (p-interaction <0.001). During the pre-Delta predominant period, the odds of SARS-CoV-2 was 66% higher (adjusted odds ratio (aOR)=1.66, 95% CI=1.43 – 1.91) among participants reporting at least one day not wearing a mask in the 10 days preceding the index date compared to those who reported no days (**Supplemental Table 3**). During the Delta-predominant period, the results were similar (aOR=1.53, 95% CI=1.23 – 1.89). This association was attenuated but remained significant during the Omicron-predominant period, where the odds of SARS-CoV-2 was 16% higher (aOR=1.16, 95% CI=1.03 – 1.32) for those reporting at least one day not wearing a mask compared with those who reported no days not wearing a mask.

## Discussion

In this community-based observational study, we found that any days not wearing a mask was associated with increased odds of SARS-CoV-2 infection after adjusting for demographics and recent known exposure. While the effect of not wearing a mask on disease transmission during non-household interactions remained significant during the entire study period, we observed a decrease in this association during the Omicron-predominant period. This variation was seen against decreasing overall mask use across the three periods.

Our findings during the pre-Delta and Delta predominant periods are consistent with previous studies suggesting that wearing a mask consistently is related to lowered odds of testing positive.^2^ To the best of our knowledge, this is the first study to extend into the Omicron predominant period and compare the protectiveness of masks.

Strengths of this study include prospectively collected data over two years on a large number of participants throughout the pandemic, and therefore is not subject to recall bias following infection. However, our findings may be limited by selection and reporting bias and may not be generalizable to other geographic regions. Other limitations include the use of self-report to determine mask use and a lack of nuance in the masking question to allow for improper use (e.g., not covering mouth or nose), type of mask (e.g., cloth, surgical or KN95/N95 mask), duration of use, and frequency and duration of interactions.

Our results suggest decreased protection for the wearer from masks during the Omicron-predominant wave. These findings may also be explained by more frequent exposures outside of one’s household later in the pandemic, increased transmissibility of the Omicron variant, high rates of vaccination and increasing population immunity during the Omicron-predominant period, and a decrease in mask wearing as guidance for vaccinated individuals evolved over time.^6,7^ While masking continues to be one of the valuable tools to decrease risk of COVID-19 infection, the level of protection for an individual wearer appears to have declined during the Omicron phase of the pandemic. Recent studies have suggested that facemasks have the potential to not only decrease odds of infection but also reduce severity of COVID-19.^8^ Future research may focus on not only odds of infection but also symptoms and severity of disease associated with mask wearing.

## Data Availability

Results of the COVID-19 CRP are being disseminated on the study website (https://www.covid19communitystudy.org/) as well as in publications and presentations in medical journals and at scientific meetings. At end of the study, the databases will be made publicly available in a de-identified manner according to CDC and applicable U.S. Federal policies.

## Declarations

## Acknowledgements

The COVID-19 Community Research Partnership gratefully acknowledges the commitment and dedication of the study participants. Programmatic, laboratory, and technical support was provided by Vysnova Partners, Inc., Javara, Inc., Oracle Corporation, LabCorp, Scanwell Health, and Neoteryx. This publication was supported by the Centers for Disease Control and Prevention (CDC) [contract #75D30120C08405] and the CARES Act of the U.S. Department of Health and Human Services (HHS) [Contract # NC DHHS GTS #49927]. Fifty percent of the current project was funded by the CDC/HHS award and fifty percent by the CARES Act/HHS award. The Partnership is listed in clinicaltrials.gov (NCT04342884). The findings and conclusions in this report are those of the authors and do not necessarily represent the official position of the Centers for Disease Control and Prevention, HHS, or the U.S. Government. A complete list of Study Sites, investigators, and staff can be found in the Appendix.

## Ethics approval

Activity was determined to meet the definition of research [45 CFR 46.102(l)] involving human subjects [45 CFR 46.102 (e)(1)] and Institutional Review Board (IRB) approval was provided by Wake Forest University.

## Consent to participate

All participants in the COVID-19 Community Research Partnership provided written consent for participation.

## Consent for publication

Not applicable. No identifying data from any individual person is contained in the manuscript.

## Author Contributions

AHT and SLE had full access to all of the data in the study and takes responsibility for the integrity of the data and the accuracy of the data analysis.

- Concept and design: All authors.
- Acquisition, analysis, or interpretation of data: All authors.
- Drafting of the manuscript: AHT, SLE
- Critical revision of the manuscript for important intellectual content: All authors.
- Statistical analysis: AHT

## Supplemental Material

**Supplemental Figure 1.**
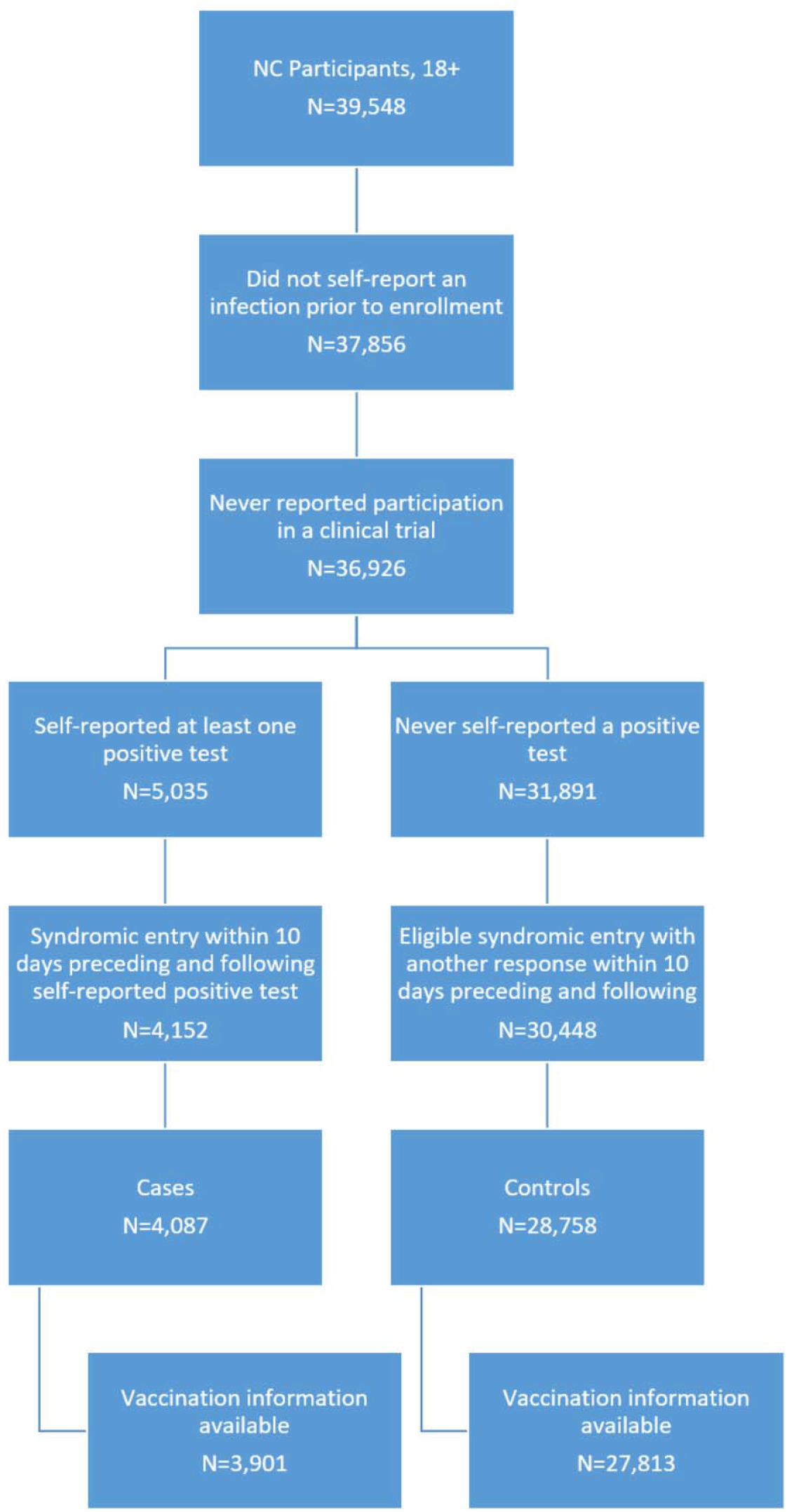
Inclusion Flow.

**Supplemental Table 1.**
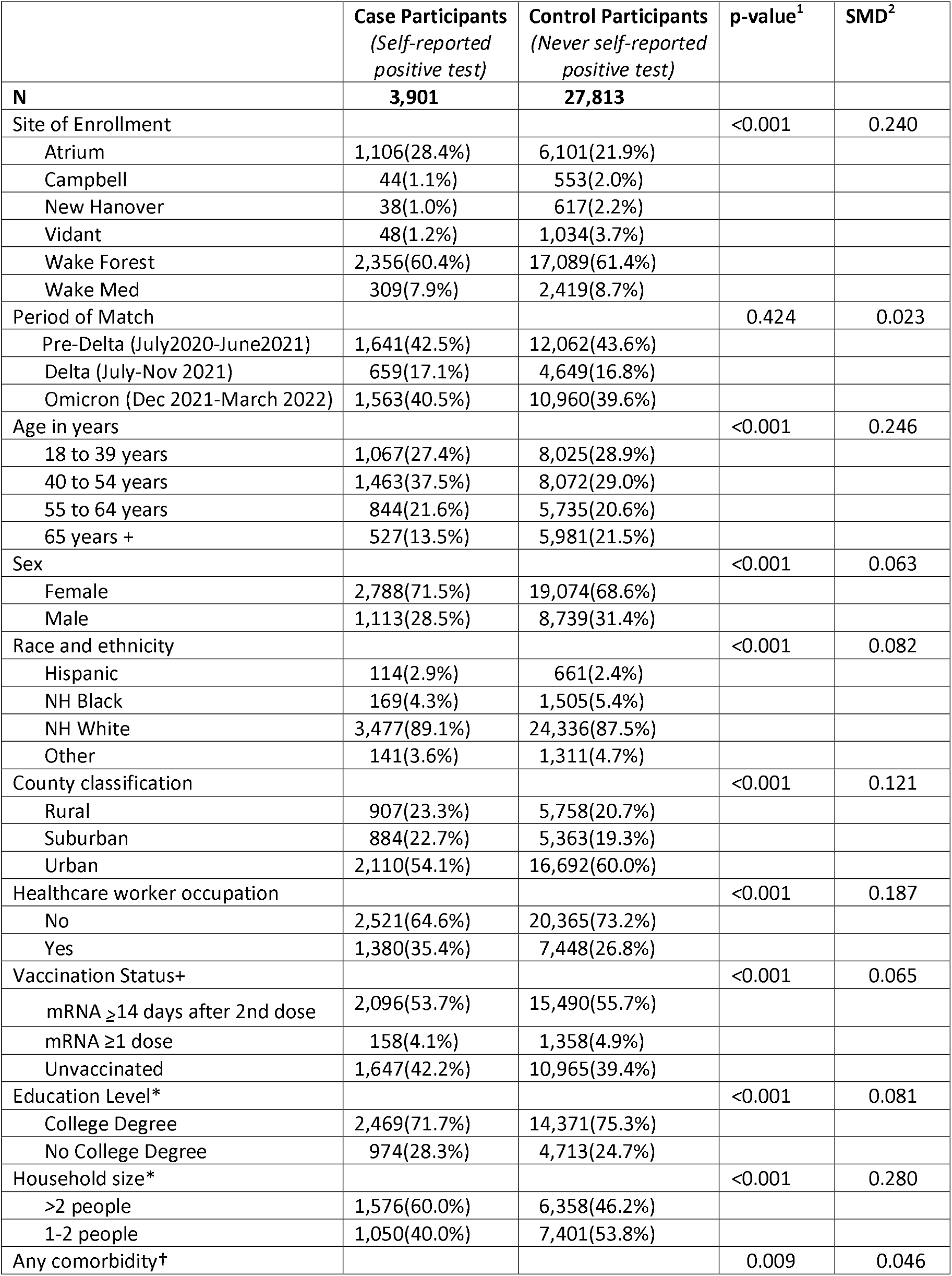
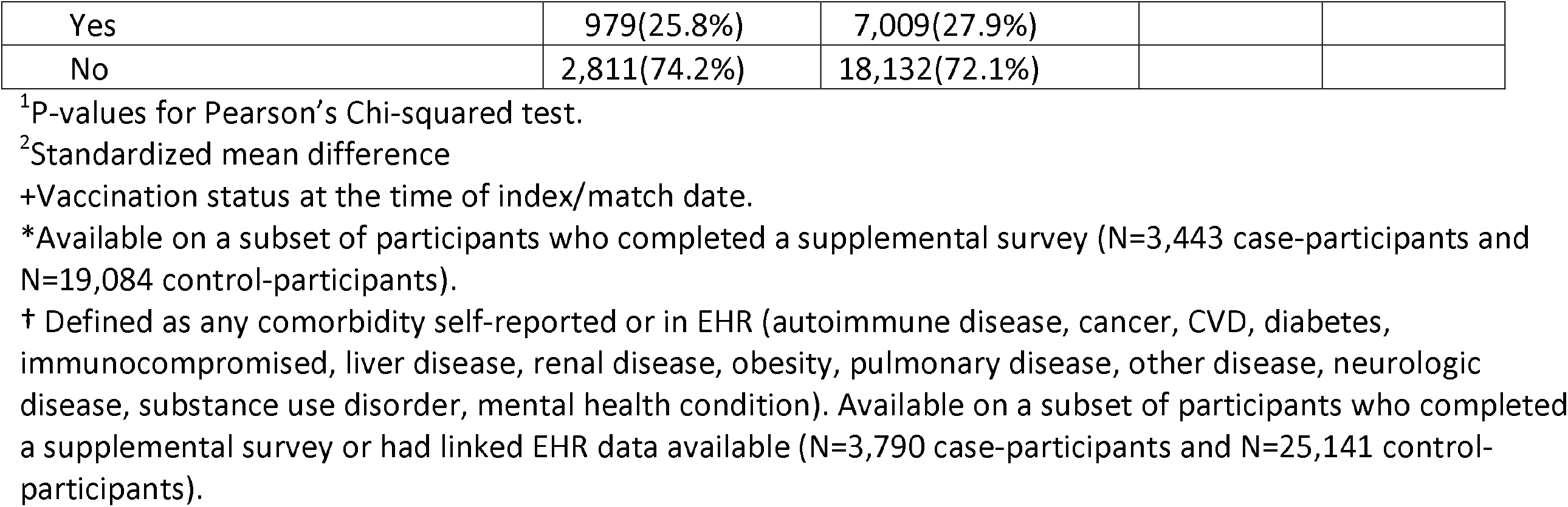
Characteristics of Participants.

|  | **Case Participants**<br>*(Self-reported positive test)* | **Control Participants**<br>*(Never self-reported positive test)* | **p-value<sup>1</sup>** | **SMD<sup>2</sup>** |
| --- | --- | --- | --- | --- |
| **N** | **3,901** | **27,813** |  |  |
| Site of Enrollment |  |  | <0.001 | 0.240 |
| Atrium | 1,106(28.4%) | 6,101(21.9%) |  |  |
| Campbell | 44(1.1%) | 553(2.0%) |  |  |
| New Hanover | 38(1.0%) | 617(2.2%) |  |  |
| Vidant | 48(1.2%) | 1,034(3.7%) |  |  |
| Wake Forest | 2,356(60.4%) | 17,089(61.4%) |  |  |
| Wake Med | 309(7.9%) | 2,419(8.7%) |  |  |
| Period of Match |  |  | 0.424 | 0.023 |
| Pre-Delta (July2020-June2021) | 1,641(42.5%) | 12,062(43.6%) |  |  |
| Delta (July-Nov 2021) | 659(17.1%) | 4,649(16.8%) |  |  |
| Omicron (Dec 2021-March 2022) | 1,563(40.5%) | 10,960(39.6%) |  |  |
| Age in years |  |  | <0.001 | 0.246 |
| 18 to 39 years | 1,067(27.4%) | 8,025(28.9%) |  |  |
| 40 to 54 years | 1,463(37.5%) | 8,072(29.0%) |  |  |
| 55 to 64 years | 844(21.6%) | 5,735(20.6%) |  |  |
| 65 years + | 527(13.5%) | 5,981(21.5%) |  |  |
| Sex |  |  | <0.001 | 0.063 |
| Female | 2,788(71.5%) | 19,074(68.6%) |  |  |
| Male | 1,113(28.5%) | 8,739(31.4%) |  |  |
| Race and ethnicity |  |  | <0.001 | 0.082 |
| Hispanic | 114(2.9%) | 661(2.4%) |  |  |
| NH Black | 169(4.3%) | 1,505(5.4%) |  |  |
| NH White | 3,477(89.1%) | 24,336(87.5%) |  |  |
| Other | 141(3.6%) | 1,311(4.7%) |  |  |
| County classification |  |  | <0.001 | 0.121 |
| Rural | 907(23.3%) | 5,758(20.7%) |  |  |
| Suburban | 884(22.7%) | 5,363(19.3%) |  |  |
| Urban | 2,110(54.1%) | 16,692(60.0%) |  |  |
| Healthcare worker occupation |  |  | <0.001 | 0.187 |
| No | 2,521(64.6%) | 20,365(73.2%) |  |  |
| Yes | 1,380(35.4%) | 7,448(26.8%) |  |  |
| Vaccination Status+ |  |  | <0.001 | 0.065 |
| mRNA ≥14 days after 2nd dose | 2,096(53.7%) | 15,490(55.7%) | | |
| mRNA ≥1 dose | 158(4.1%) | 1,358(4.9%) | | |
| Unvaccinated | 1,647(42.2%) | 10,965(39.4%) |  |  |
| Education Level* |  |  | <0.001 | 0.081 |
| College Degree | 2,469(71.7%) | 14,371(75.3%) |  |  |
| No College Degree | 974(28.3%) | 4,713(24.7%) |  |  |
| Household size* |  |  | <0.001 | 0.280 |
| >2 people | 1,576(60.0%) | 6,358(46.2%) |  |  |
| 1-2 people | 1,050(40.0%) | 7,401(53.8%) |  |  |
| Any comorbidity† |  |  | 0.009 | 0.046 |
| Yes | 979(25.8%) | 7,009(27.9%) |  |  |
| No | 2,811(74.2%) | 18,132(72.1%) |  |  |

<sup>1</sup>P-values for Pearson's Chi-squared test.
<sup>2</sup>Standardized mean difference
+Vaccination status at the time of index/match date.
\*Available on a subset of participants who completed a supplemental survey (N=3,443 case-participants and N=19,084 control-participants).
† Defined as any comorbidity self-reported or in EHR (autoimmune disease, cancer, CVD, diabetes, immunocompromised, liver disease, renal disease, obesity, pulmonary disease, other disease, neurologic disease, substance use disorder, mental health condition). Available on a subset of participants who completed a supplemental survey or had linked EHR data available (N=3,790 case-participants and N=25,141 control-participants).

**Supplemental Table 2.** Self-Reported Behavioral Characteristics, 10 days Preceding Match Date.

|  | **Case Participants**<br>*(Self-reported positive test)* | **Control Participants**<br>*(Never self-reported positive test)* | **p-value<sup>1</sup>** | **SMD<sup>2</sup>** |
| --- | --- | --- | --- | --- |
| **N** | **3,901** | **27,813** |  |  |
| Daily Survey Response Rate | 0.731±0.297 | 0.653±0.327 | <0.001 | 0.250 |
| Any days interaction without mask* |  |  | <0.001 | 0.126 |
| No (no days) | 2,244(57.5%) | 17,706(63.7%) |  |  |
| Yes (at least one day) | 1,657(42.5%) | 10,107(36.3%) |  |  |
| Proportion days interaction without mask* | 0.18±0.28 | 0.16±0.28 | <0.001 | 0.086 |
| Any known exposure |  |  | <0.001 | 1.100 |
| No | 1,789(45.9%) | 25,227(90.7%) |  |  |
| Yes | 2,112(54.1%) | 2,586(9.3%) |  |  |
| Any days reporting at least one symptoms |  |  | <0.001 | 1.312 |
| No | 703(18.0%) | 20,174(72.7%) |  |  |
| Yes | 3,192(82.0%) | 7,593(27.3%) |  |  |
| Any days reporting 3+ symptoms |  |  | <0.001 | 1.277 |
| No | 1,642(42.1%) | 25,747(92.6%) |  |  |
| Yes | 2,259(57.9%) | 2,066(7.4%) |  |  |
| Sought treatment |  |  | <0.001 | 0.542 |
| No | 3,086(79.1%) | 26,780(96.3%) |  |  |
| Yes | 815(20.9%) | 1,033(3.7%) |  |  |

<sup>1</sup>P-values for Pearson's Chi-squared test for categorical variables and Welch's two-sample t-test for continuous variables.
<sup>2</sup>Standardized mean difference
\*Participants who responded "No" to the question: "In the last 24 hours, have you worn a face mask or face covering every time you interacted with others (not in your household) within a distance of less than 6 feet?"

**Supplemental Table 3.**
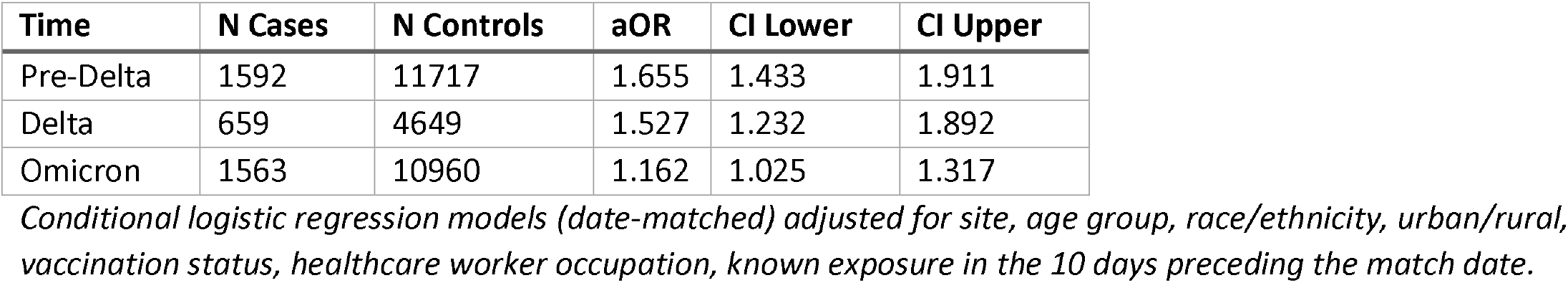
Effect of Self-Reporting Any Days Not Wearing a Mask when Interacting with others Outside Household on Self-Reported SARS-CoV-2 Infection by Period

## Authorship Appendix

**The COVID-19 Research Group** (*Site Principal Investigator)

### Wake Forest School of Medicine

Thomas F Wierzba PhD, MPH, MS*, John Walton Sanders, MD, MPH, David Herrington, MD, MHS, Mark A. Espeland, PhD, MA, John Williamson, PharmD, Morgana Mongraw-Chaffin, PhD, MPH, Alain Bertoni, MD, MPH, Martha A. Alexander-Miller, PhD, Paola Castri, MD, PhD, Allison Mathews, PhD, MA, Iqra Munawar, MS, Austin Lyles Seals, MS, Brian Ostasiewski, Christine Ann Pittman Ballard, MPH, Metin Gurcan, PhD, MS, Alexander Ivanov, MD, Giselle Melendez Zapata, MD, Marlena Westcott, PhD, Karen Blinson, Laura Blinson, Mark Mistysyn, Donna Davis, Lynda Doomy, Perrin Henderson, MS, Alicia Jessup, Kimberly Lane, Beverly Levine, PhD, Jessica McCanless, MS, Sharon McDaniel, Kathryn Melius, MS, Christine O’Neill, Angelina Pack, RN, Ritu Rathee, RN, Scott Rushing, Jennifer Sheets, Sandra Soots, RN, Michele Wall, Samantha Wheeler, John White, Lisa Wilkerson, Rebekah Wilson, Kenneth Wilson, Deb Burcombe, Georgia Saylor, Megan Lunn, Karina Ordonez, Ashley O’Steen, MS, Leigh Wagner.

### Atrium Health

Michael S. Runyon MD, MPH*, Lewis H. McCurdy MD*, Michael A. Gibbs, MD, Yhenneko J. Taylor, PhD, Lydia Calamari, MD, Hazel Tapp, PhD, Amina Ahmed, MD, Michael Brennan, DDS, Lindsay Munn, PhD RN, Keerti L. Dantuluri, MD, Timothy Hetherington, MS, Lauren C. Lu, Connell Dunn, Melanie Hogg, MS, CCRA, Andrea Price, Marina Leonidas, Melinda Manning, Whitney Rossman, MS, Frank X. Gohs, MS, Anna Harris, MPH, Jennifer S. Priem, PhD, MA, Pilar Tochiki, Nicole Wellinsky, Crystal Silva, Tom Ludden PhD, Jackeline Hernandez, MD, Kennisha Spencer, Laura McAlister.

### MedStar Health Research Institute

William Weintraub MD*, Kristen Miller, DrPH, CPPS*, Chris Washington, Allison Moses, Sarahfaye Dolman, Julissa Zelaya-Portillo, John Erkus, Joseph Blumenthal, Ronald E. Romero Barrientos, Sonita Bennett, Shrenik Shah, Shrey Mathur, Christian Boxley, Paul Kolm, PhD, Ella Franklin, Naheed Ahmed, Moira Larsen.

### Tulane

Richard Oberhelman MD*, Joseph Keating PhD*, Patricia Kissinger, PhD, John Schieffelin, MD, Joshua Yukich, PhD, Andrew Beron, MPH, Johanna Teigen, MPH.

### University of Maryland School of Medicine

Karen Kotloff MD*, Wilbur H. Chen MD, MS*, DeAnna Friedman-Klabanoff, MD, Andrea A. Berry, MD, Helen Powell, PhD, Lynnee Roane, MS, RN, Reva Datar, MPH, Colleen Reilly.

### University of Mississippi

Adolfo Correa MD, PhD*, Bhagyashri Navalkele, MD, Yuan-I Min, PhD, Alexandra Castillo, MPH, Lori Ward, PhD, MS, Robert P. Santos, MD, MSCS, Pramod Anugu, Yan Gao, MPH, Jason Green, Ramona Sandlin, RHIA, Donald Moore, MS, Lemichal Drake, Dorothy Horton, RN, Kendra L. Johnson, MPH, Michael Stover.

### Wake Med Health and Hospitals

William H. Lagarde MD*, LaMonica Daniel, BSCR.

### New Hanover

Patrick D. Maguire MD*, Charin L. Hanlon, MD, Lynette McFayden, MSN, CCRP, Isaura Rigo, MD, Kelli Hines, BS, Lindsay Smith, BA, Monique Harris, CCRP, Belinda Lissor, AAS, CCRP, Vivian Cook, MA, MPH, Maddy Eversole, BS, Terry Herrin, BS, Dennis Murphy, RN, Lauren Kinney, BS, Polly Diehl, MS, RHIA, Nicholas Abromitis, BS, Tina St. Pierre, BS, Bill Heckman, Denise Evans, Julian March, BA, Ben Whitlock, CPA, MSA, Wendy Moore, BS, AAS, Sarah Arthur, MSW, LCSW, Joseph Conway.

### Vidant Health

Thomas R. Gallaher MD*, Mathew Johanson, MHA, CHFP, Sawyer Brown, MHA, Tina Dixon, MPA, Martha Reavis, Shakira Henderson, PhD, DNP, MS, MPH, Michael Zimmer, PhD, Danielle Oliver, Kasheta Jackson, DNP, RN, Monica Menon, MHA, Brandon Bishop, MHA, Rachel Roeth, MHA.

### Campbell University School of Osteopathic Medicine

Robin King-Thiele DO*, Terri S. Hamrick PhD*, Abdalla Ihmeidan, MHA, Amy Hinkelman, PhD, Chika Okafor, MD (Cape Fear Valley Medical Center), Regina B. Bray Brown, MD, Amber Brewster, MD, Danius Bouyi, DO, Katrina Lamont, MD, Kazumi Yoshinaga, DO, (Harnett Health System), Poornima Vinod, MD, A. Suman Peela, MD, Giera Denbel, MD, Jason Lo, MD, Mariam Mayet-Khan, DO, Akash Mittal, DO, Reena Motwani, MD, Mohamed Raafat, MD (Southeastern Health System), Evan Schultz, DO, Aderson Joseph, MD, Aalok Parkeh, DO, Dhara Patel, MD, Babar Afridi, DO (Cumberland County Hospital System, Cape Fear Valley).

### George Washington University Data Coordinating Center

Diane Uschner PhD*, Sharon L. Edelstein, ScM, Michele Santacatterina, PhD, Greg Strylewicz, PhD, Brian Burke, MS, Mihili Gunaratne, MPH, Meghan Turney, MA, Shirley Qin Zhou, MS, Ashley H Tjaden, MPH, Lida Fette, MS, Asare Buahin, Matthew Bott, Sophia Graziani, Ashvi Soni, MS.

### George Washington University Mores Lab

Christopher Mores, PhD, Abigail Porzucek, MS.

### Oracle Corporation

Rebecca Laborde, Pranav Acharya.

### Sneez LLC

Lucy Guill, MBA, Danielle Lamphier, MBA, Anna Schaefer, MSM, William M. Satterwhite, JD, MD.

### Vysnova Partners

Anne McKeague, PhD, Johnathan Ward, MS, Diana P. Naranjo, MA, Nana Darko, MPH, Kimberly Castellon, BS, Ryan Brink, MSCM, Haris Shehzad, MS, Derek Kuprianov, Douglas McGlasson, MBA, Devin Hayes, BS, Sierra Edwards, MS, Stephane Daphnis, MBA, Britnee Todd, BS.

### Javara Inc

Atira Goodwin.

### External Advisory Council

Ruth Berkelman, MD, Emory, Kimberly Hanson, MD, U of Utah, Scott Zeger, PhD, Johns Hopkins, Cavan Reilly, PhD, U. of Minnesota, Kathy Edwards, MD, Vanderbilt, Helene Gayle, MD MPH, Chicago Community Trust, Stephen Redd.

